# Androgen Deprivation Therapy (ADT) and Radiotherapy (RT) with Imaging Evaluation Longitudinally (ARIEL) trial: protocol, early results, and implications of neoadjuvant ADT for focal RT boost in prostate cancer

**DOI:** 10.64898/2026.04.22.26351215

**Authors:** Yuze Song, Mariluz Rojo Domingo, Lily Nguyen, Christopher C Conlin, Nitin Dhillon, Son Do, Anna Dornisch, Michael E Hahn, Roshan Karunamuni, Joshua Kim, Kang-Lung Lee, Jasmine Liu, Rana R McKay, Loren K Mell, Arno J Mundt, Rakesh Patel, Edmund M Qiao, Brent Rose, Rhea Rupareliya, Hadley Schaub, Armin Schwartzman, Tyler Stewart, Anders M Dale, Tyler M Seibert ARIEL investigators

## Abstract

**Background:** Men with aggressive, localized prostate cancer (PC) undergo definitive radiotherapy (RT) with androgen deprivation therapy (ADT). The prospective, phase II ARIEL trial evaluates a quantitative MRI biomarker, Restriction Spectrum Imaging restriction score (RSIrs), at three time points (before treatment, after ADT and after RT) for treatment response assessment. RSIrs highlights intracellular restricted diffusion and is correlated with high-grade PC.

**Design:** Participants are men with unfavorable-intermediate-risk or high-risk localized PC undergoing definitive RT with neoadjuvant and concurrent ADT, and MRI-RSI acquisitions at three time points: before therapy, after neoadjuvant ADT but before RT, and after RT. The primary aim is to evaluate performance of RSIrs for identifying patients who will experience early biochemical recurrence. Change in RSIrs within visible tumors after ADT and RT is the primary independent variable.

**Results:** 97 patients met inclusion criteria and received ≥1 MRI. On central review, visible PI-RADS lesions were identified in 88 patients: 80 patients had one lesion, and 8 patients had two lesions. After neoadjuvant ADT, 40% of lesions were not clearly visible. Those still visible had shrank by median 55.8% (IQR: 42.8-69.0%), much more than the prostate volume decrease of 21.5% (11.9-31.6%). RSIrs maximum within visible lesions decreased from mean 329 (SD:185) pre-ADT to 209 (SD:125) pre-RT (*p*<0.01), and to 107 (SD:61) post-RT (*p*<0.01). Conventional apparent diffusion coefficient (ADC) changes were less consistent. Follow-up is ongoing to assess whether imaging response is related to future recurrence risk.

**Conclusion:** ARIEL has completed accrual and preliminary results demonstrate changes in RSIrs after treatment, which may indicate tumor response. Primary results will be presented when the primary endpoint is reached. With neoadjuvant ADT, both pre- and post-ADT MRI are likely necessary for accurate focal RT boost targeting. Concurrent commencement of ADT and RT simplifies workflows and facilitates accurate gross tumor volume delineation.

## Background

Men with localized prostate cancer (PC) with higher risk features may develop incurable metastatic disease and benefit from definitive radiation therapy (RT) combined with androgen deprivation therapy (ADT)^1^. While this treatment is highly effective overall, some patients with adverse PC will experience recurrence. Prostate-specific antigen (PSA) is the standard metric for detecting recurrence, but PSA surveillance has significant drawbacks for post-treatment monitoring. PSA values may take years to reach their post-treatment nadir. Moreover, PSA is suppressed during concurrent ADT, limiting its immediate diagnostic utility. Early detection of treatment failure may be crucial for implementing salvage therapies while they remain potentially curative. Likewise, early confirmation of favorable response to treatment might facilitate shortening of treatment duration to improve quality of life. Therefore, there is a need for more sophisticated biomarkers to reliably predict treatment outcomes and guide personalized therapeutic decisions in high-risk populations. Advanced imaging techniques may provide more timely and accurate assessment of treatment response.

The most useful magnetic resonance imaging (MRI) modality for PC detection is diffusion-weighted imaging (DWI). However, the standard DWI biomarker (the apparent diffusion coefficient [ADC]) combines signal from intracellular and extracellular microcompartments into a single value that cannot account for the many varied changes that occur in prostate tissue after ADT and RT. Restriction Spectrum Imaging (RSI) is a multi-compartment model of DWI that separates tissue microenvironments based on their diffusion characteristics (restricted intracellular, hindered extracellular, free diffusion and flow^2–4^). The RSI restriction score (RSIrs) is an RSI-based quantitative imaging biomarker that highlights regions of the prostate with intracellular restricted diffusion to distinguish between cancerous and non-cancerous prostatic tissue^5^. RSIrs is also more reproducible than ADC^6–8^. We hypothesized that quantitative post-treatment changes in RSIrs could serve as an indicator of therapeutic response, with persistently elevated values serving as a predictive biomarker for increased risk of recurrence. Because RSIrs is specifically designed to emphasize intracellular restricted diffusion—rather than averaging all types of diffusion of with a single parameter like ADC—it may offer improved sensitivity and specificity for identifying residual or recurrent disease after therapy.

Longitudinal MRI may also inform how to manage logistical challenges of combining focal RT boost strategies when coordinated with ADT, particularly regarding optimal timing and sequencing of treatments^9–11^. When ADT is initiated before RT, delivering targeted boost doses is complicated by changes in prostate size and shape, as well as decreased conspicuity of tumors on conventional MRI, which can compromise accurate target delineation. MRI monitoring throughout the treatment process on the ARIEL trial could provide valuable, quantitative insights into prostate and tumor changes over time, including treatment-induced alterations in diffusion characteristics. Such longitudinal imaging data could inform the optimal timing of ADT initiation relative to RT planning, balancing adequate tumor visualization for precise targeting with the goal of maximizing therapeutic efficacy.

### Rationale for trial

Serum PSA levels are routinely used to monitor response to androgen deprivation therapy (ADT) and radiotherapy (RT). Although PSA decreases in most patients following treatment, a subset will ultimately experience disease recurrence. Early PSA kinetics alone are not reliably predictive of subsequent treatment failure. There is a need for better biomarkers to predict earlier recurrence and to guide personalized post-treatment oncologic care. The ADT and Radiotherapy with Imaging Evaluation Longitudinally (ARIEL) trial is evaluating a novel imaging biomarker, RSIrs, for identifying participants who will experience early biochemical recurrence (i.e., within 3 years of post-treatment). A secondary endpoint is failure to reach a PSA nadir <0.5 ng/mL within 18 months of RT, indicating a poorer prognosis and higher risk of recurrence. We hypothesize that the change in RSIrs within visible prostate tumors from MRI #1 (acquired before any treatment) to MRI #3 (acquired after ADT and RT) will be associated with early biochemical recurrence. In this paper, we describe the trial design and, as both accrual and imaging are complete, present the participant characteristics and preliminary findings, including observed RSIrs changes in intraprostatic lesions.

### Design

#### Overview and Schema

ARIEL is a prospective, phase II, single-arm, imaging biomarker trial designed to evaluate change in RSIrs for association with risk of biochemical recurrence. The study was approved by the Institutional Review Board, and all participants provided informed consent. Participants are men with unfavorable-intermediate-risk or high-risk localized PC undergoing definitive RT with neoadjuvant and concurrent ADT. Participants undergo MRI with RSI acquisitions at three time points: before therapy (MRI #1), after neoadjuvant ADT but before RT (MRI #2), and after RT (MRI #3). The protocol allowed for omitting MRI #2 because of logistical considerations for starting RT, if necessary. Treatment response will be primarily assessed by absence of biochemical recurrence. The primary objective is to evaluate the performance of RSIrs for identifying patients who will experience early biochemical recurrence (i.e., within 3 years of completing RT). Change in RSIrs within visible tumors from MRI #1 to MRI #3 will be the primary independent variable. A summary of the trial schema is illustrated in Figure 1.

**Figure 1.**
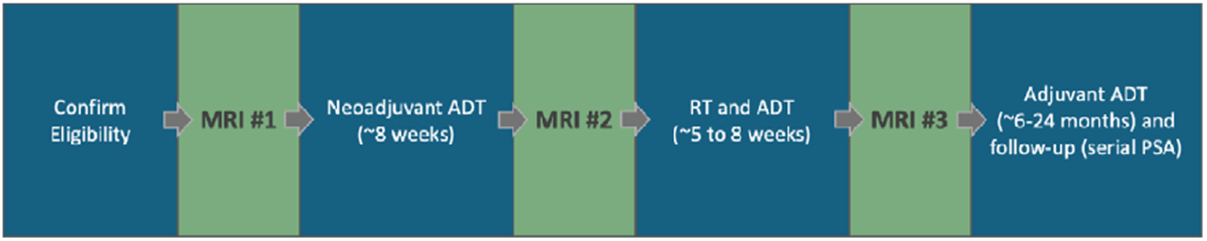
ARIEL trial schema. Potential participants were screened based on the inclusion and exclusion criteria, and provided informed consent to participate in the study. Baseline assessments were performed for each participant before receiving three serial MRIs with diffusion-based Restriction Spectrum Imaging (RSI) acquisitions: MRI #1 took place before starting androgen deprivation therapy (ADT) and radiotherapy (RT), MRI #2 after starting ADT but before starting RT, and MRI #3 was after receiving both ADT and RT. After MRI #3, adjuvant ADT was administered. For follow-up, prostate-specific antigen (PSA) levels are routinely measured for at least 60 months after all MRIs are acquired.

#### Inclusion and exclusion criteria

Eligible participants were men aged ≥18 years with histologically confirmed PC classified as either high-risk (PSA ≥20 ng/mL, cT3-T4 stage, or Grade Group [GG] ≥4) or unfavorable intermediate-risk (≥50% positive biopsy cores, GG 3, or two intermediate risk factors including GG 2, cT2b-T2c stage, and PSA 10-20 ng/mL). Participants planned to undergo standard-of-care definitive RT with neoadjuvant and concurrent ADT, had good general health (ECOG performance status 0-2), were willing to comply with all study procedures and follow-up requirements, and provided informed consent to participate in the trial. Patients were excluded if they had prior RT to the pelvis, previous PC treatment (cryotherapy, HIFU, prostatectomy), hip prostheses (they can introduce severe image artifacts), or MRI contraindications per institutional guidelines. Additional exclusion criteria included clear evidence of nodal or distant metastasis on conventional staging imaging, active malignancy that could impact survival or standard care delivery, or any medical condition, therapy, or laboratory abnormality that could confound study results, interfere with full participation, or compromise patient safety.

#### Study treatment and imaging

Dose and fractionation for RT and ADT were delivered per standard clinical practice. Standard of care therapy included ADT with a luteinizing hormone-releasing hormone agonist or antagonist. Duration of ADT was not specified on the trial but expected to be typically 4-6 months for unfavorable-intermediate-risk PC or 18-24 months for high-risk PC. Addition of androgen receptor blockers or androgen receptor pathway inhibitors was allowed at the discretion of treating physicians. ADT began 2-3 months before RT, which was delivered by an external beam technique (with photons, protons or both) with standard fractionation or moderate hypofractionation, delivered 5 days per week for 4-9 weeks. Treatment of regional lymph node regions was added on a case-by-case basis per judgment of the treating physician. Per clinical routine, the timing and duration of any concomitant PC therapy falling within the routine described above (or any individual variations) were recorded during screening and at every clinic visit. As this was an observational study, all treatment decisions were pragmatic and reflective of real-world variability in prostate RT practice.

Study imaging consisted of three serial MRI scans of the prostate with multi-*b*-value diffusion RSI acquisitions. No intravenous contrast was administered (unless the scan coincided with a clinical multiparametric MRI exam), and no endorectal coil was used. All institutional safety procedures, protocols, and policies were applied, per clinical routine. Imaging was acquired on 3T GE Healthcare scanners at a single center. Each MRI included axial *T*_*2*_-weighted and RSI diffusion series, with imaging parameters that matched those used in multiparametric prostate MRI for clinical care (Supplementary Table 1).

RSI data were processed to correct for noise, gradient nonlinearities and eddy currents using custom MATLAB scripts (MathWorks, USA). RSIrs was then computed from the corrected image data. RSI models the diffusion signal as four micro-compartments (Supplementary Table 2), and the RSIrs biomarker reflects normalized intracellular signal, highest in regions of restricted diffusion typical of clinically significant PC. To assess repeatability and reproducibility, the RSI acquisition was repeated once at minimum echo time and once more at a longer echo time (allowing estimation of quantitative 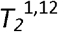). After MRI #1, ADT was initiated and was administered for 2-3 months before MRI #2 (minimum 5 weeks). MRI#3 was performed after completing RT, ideally within 4 weeks. After MRI #3, participants received adjuvant ADT to complete the full prescribed duration. Follow-up in this observational trial was per clinical routine with serial PSA tests.

#### Study objectives and endpoints

The primary endpoint is the prognostic accuracy of RSIrs in identifying patients with biochemical recurrence (PSA≥2 ng/mL greater than nadir) within 3 years of RT completion. Accuracy will be expressed as sensitivity and specificity for all possible cut-points, presented with receiver operating characteristic (ROC) curves, and summarized as the area under the ROC curve (AUC). The clinical motivation for this endpoint is that biochemical recurrence signals treatment failure and a higher risk of death from metastasis. Adding effective therapies at initial treatment may improve survival and long-term quality of life, so early identification of patients at high risk is relevant. Conversely, identifying favorable responders might inform future trials testing shorter duration of ADT. The secondary endpoint is the prognostic accuracy of RSIrs in identifying patients who have a PSA nadir ≤0.5 ng/mL during the first 18 months after completing RT, a favorable prognostic indicator associated with improved progression-free survival^13^.

#### Data collection and follow-up

The study duration will be 6 years, and the participation duration was 5 months. All study imaging was collected in a single center, although participants were allowed to receive routine treatment at other sites. Follow-up was per clinical routine, and data were obtained via chart review.

#### Statistical planning for pre-specified outcomes

We will analyze data from all participants according to the intention-to-treat principle. For the primary analysis, a logistic regression model of biochemical recurrence within 3 years will be fit to the change in RSIrs from MRI #1 to MRI #3 and relevant covariates (e.g., age, duration of ADT). Continuous variables will be splined, and only main effects will be considered. We will estimate the non-parametric ROC and its AUC of the change in RSIrs using biochemical recurrence within 3 years as the reference standard. Cross-validation will be performed, and we will report 95% bootstrap confidence intervals (CI) for AUCs. If the lower bound of the AUC 95% confidence interval exceeds 0.55, RSIrs will be considered to demonstrate discriminatory performance for identifying participants who recur within 3 years post-treatment. In that case, a Cox model for predicting recurrence time will be fit and the hazard ratios for RSI will be used to describe the adjusted associations supported by the data between RSIrs and recurrence time. Time-dependent ROC curves will be constructed to leverage the actual time-to-recurrence. From this model, we will graph the AUC as a function of the time-to-recurrence to examine the predictive accuracy of RSIrs for early and late recurrent events. A secondary analysis will repeat the above approach but will evaluate performance of RSIrs before and during therapy as a biomarker to identify patients who will fail to have PSA nadir <0.5 ng/mL within 18 months post-RT. Exploratory analyses will repeat the above but use change in RSIrs from MRI#1 to MRI#2 as the independent variable (i.e., response to neoadjuvant ADT, prior to starting RT).

### Methods for Preliminary Analysis

To investigate changes to the prostate and tumor appearance with treatment, automatic segmentation of the prostate was performed for each time point (MRI #1, MRI #2, MRI #3) using a validated AI model^14–16^ to obtain preliminary volumes. These prostate contours were then reviewed centrally by a single genitourinary radiologist (5 years of experience). All auto-segmentations underwent centralized review by a single board-certified genitourinary radiologist with expertise in prostate MRI. Each case was classified as acceptable or unacceptable, and minor manual corrections were performed when necessary. The radiologist also drew the intraprostatic lesions using the Prostate Imaging Reporting and Data System (PI-RADS v2.1)^17^. We measured prostate and lesion volume changes after neoadjuvant ADT, and after ADT+RT. We also calculated the percentage of lesions that remained clearly visible (as judged by the central radiology review) after ADT and after ADT+RT. For lesions that remained MRI-visible, we computed the change in size, normalized by prostate volume change, to determine whether the cancerous areas shrank relatively more or less than the prostate.

Finally, we computed quantitative changes in maximum RSIrs and mean ADC for MRI #1 vs. MRI #2, and for MRI #1 vs. MRI #3. Maximum RSIrs and mean ADC were obtained for each patient within the entire prostate and for the most suspicious intraprostatic lesion (highest PI-RADS, then largest lesion if PI-RADS scores were tied). For each participant, we obtained the RSIrs and ADC values at each time point to observe the change in values over time and used *t*-tests to identify significant changes in quantitative values. We used violin plots to illustrate overall changes at the cohort-level and dot plots to show the individual variability in biomarker changes from participant to participant.

### Preliminary Results

Of 123 patients, 26 patients were excluded because they no longer wanted to participate in the study, started ADT too early before MRI #1 was acquired, or declined ADT and/or RT (Supplementary Figure 1). The demographic and clinical characteristics for the participants are summarized in Table 1. The ADT agents used were leuprolide, degarelix, and relugolix. RT was delivered via volumetric modulated arc therapy (VMAT), stereotactic body radiation therapy (SBRT) or intensity modulated proton therapy (IMPT).

Of the 97 patients that met the inclusion criteria and received baseline MRI #1, 94 patients also received MRI #3, and 79 patients also received MRI #2. On central radiology review, visible PI-RADS lesions were identified in 88 of the 97 included patients, with a total of 96 lesions contoured for MRI #1: 80 patients had one PI-RADS lesion, and 8 patients had two PI-RADS lesions. Among the 94 patients that had MRI #3 (acquired after both ADT and RT), 85 patients had visible lesions, with a total of 93 lesions (8 of the 93 lesions [9%] remained clearly visible after RT). Among the 79 patients that had MRI #2 (acquired after ADT), 72 patients had visible lesions, with a total of 78 lesions (46 of the 78 lesions [60%] remained clearly visible after ADT). Examples of RSI maps and PI-RADS lesions overlaid on T2W anatomical images before and after treatment are shown in Figure 2.

**Figure 2.**
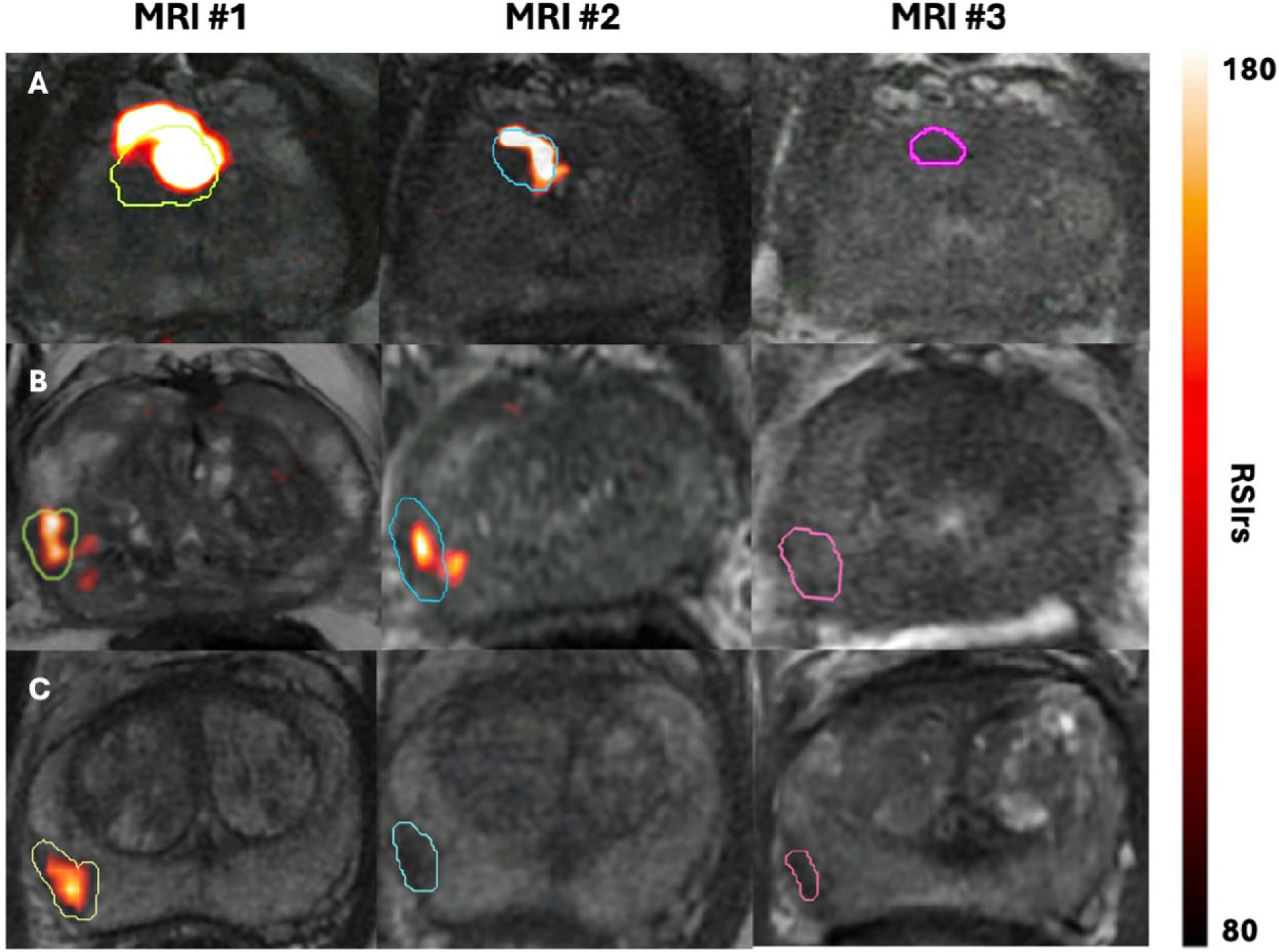
Restriction Spectrum Imaging (RSI) maps and Prostate Imaging Reporting and Data System (PI-RADS) lesions overlaid on T2-weighted anatomical images at three timepoints: pre-treatment (MRI #1), after ADT but before RT (MRI #2), and after ADT and RT (MRI #3). The top row (patient A) corresponds to serial MRIs for a patient with grade group 2 prostate cancer, the middle row (B) for a patient with grade group 4 disease, and the bottom row (C) for a patient with grade group 3 cancer. Changes in the intensity of RSI maps and the size of the PI-RADS lesions drawn by the radiologist are observed.

Prostate and lesion volume changed after ADT and RT (Table 2, Supplementary Figure 2). The median percentage decrease in prostate volume was 21.5% (IQR: 11.9-31.6%) from MRI #1 to MRI #2, and 32.3% (IQR: 24.8-41.4%) from MRI #1 to MRI #3. The median percentage decrease in lesion volume was 55.8% (IQR: 42.8-69.0) from MRI #1 to MRI #2, and 72.8% (IQR: 57.8-80.0%) from MRI #1 to MRI #3.

The cohort’s maximum RSIrs and mean ADC values within visible lesions at each timepoint are shown in Figure 3. Before ADT, the mean RSIrs maximum was 329 (standard deviation [SD]: 185), and it significantly decreased to 209 (SD: 125) after ADT (*p*<0.01), then even more after RT, to a mean value of 107 (SD 61; *p*<0.01). Mean lesion ADC significantly increased from MRI #1 to MRI #3 (914×10^-6^ mm^2^/s vs. 1064×10^-6^ mm^2^/s, *p*<0.01), but was not significantly different between MRI #1 and MRI #2 (914×10^-6^ mm^2^/s vs. 935×10^-6^ mm^2^/s, *p*=0.67). Violin plots showing the cohort’s changes in maximum RSIrs and mean ADC in the entire prostate are shown in Supplementary Figure 3.

**Figure 3.**
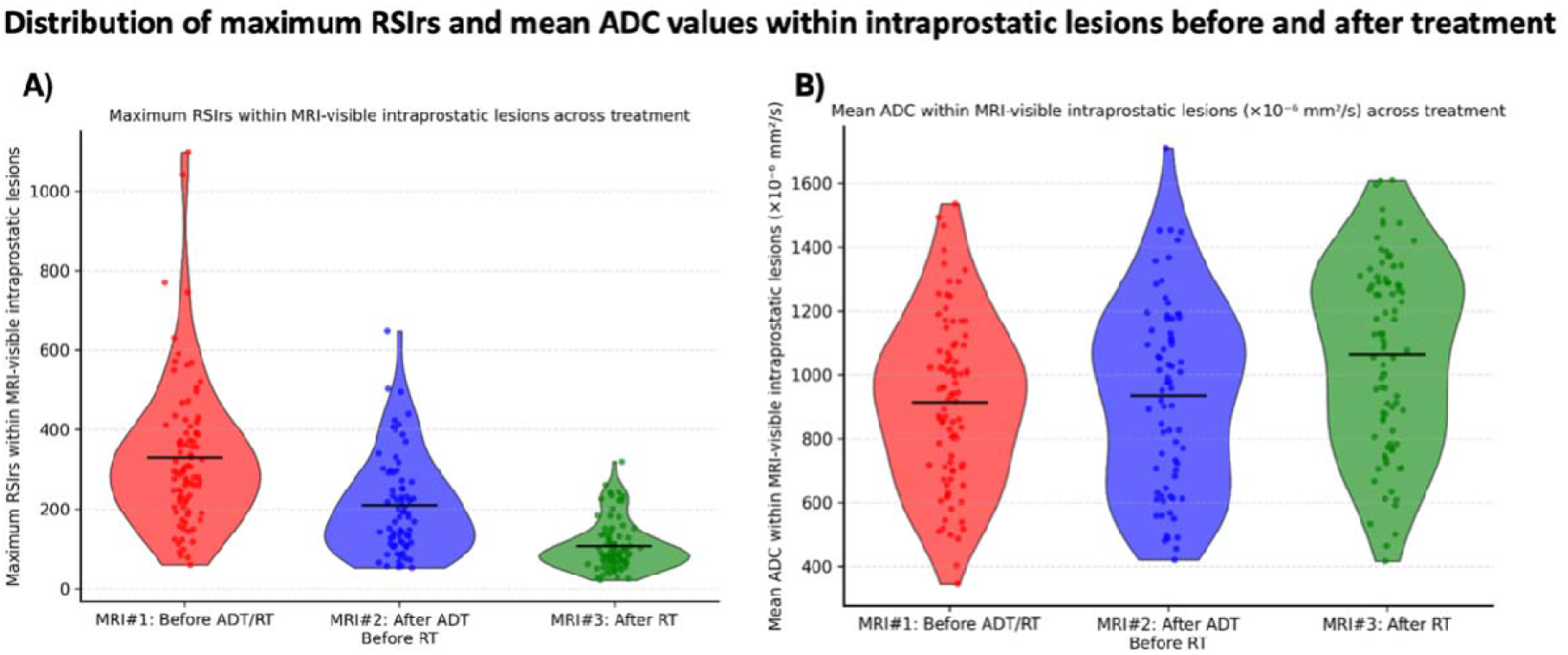
Maximum Restriction Spectrum Imaging restriction score (RSIrs) and mean apparent diffusion coefficient (ADC) values within the MRI-visible lesions in the prostate for patients included in the study. MRI #1 was acquired before ADT and RT, MRI #2 after ADT but before RT, and MRI #3 was after RT. RSIrs values decreased after ADT, and also after RT. The increase in mean ADC values after treatment is more moderate.

Individual changes in maximum RSIrs and mean ADC within MRI-visible lesions are shown in Figure 4 (and for maximum within the entire prostate in Supplementary Figure 4). Prostate auto-segmentation worked well in the pre- and post-treatment settings. The radiologist classified all prostate auto-segmentations as acceptable; no prostate auto-segmentations required significant modifications from the radiologist to obtain the final prostate contour (median Dice Coefficient = 0.89 [IQR: 0.86-0.91]).

**Figure 4.**
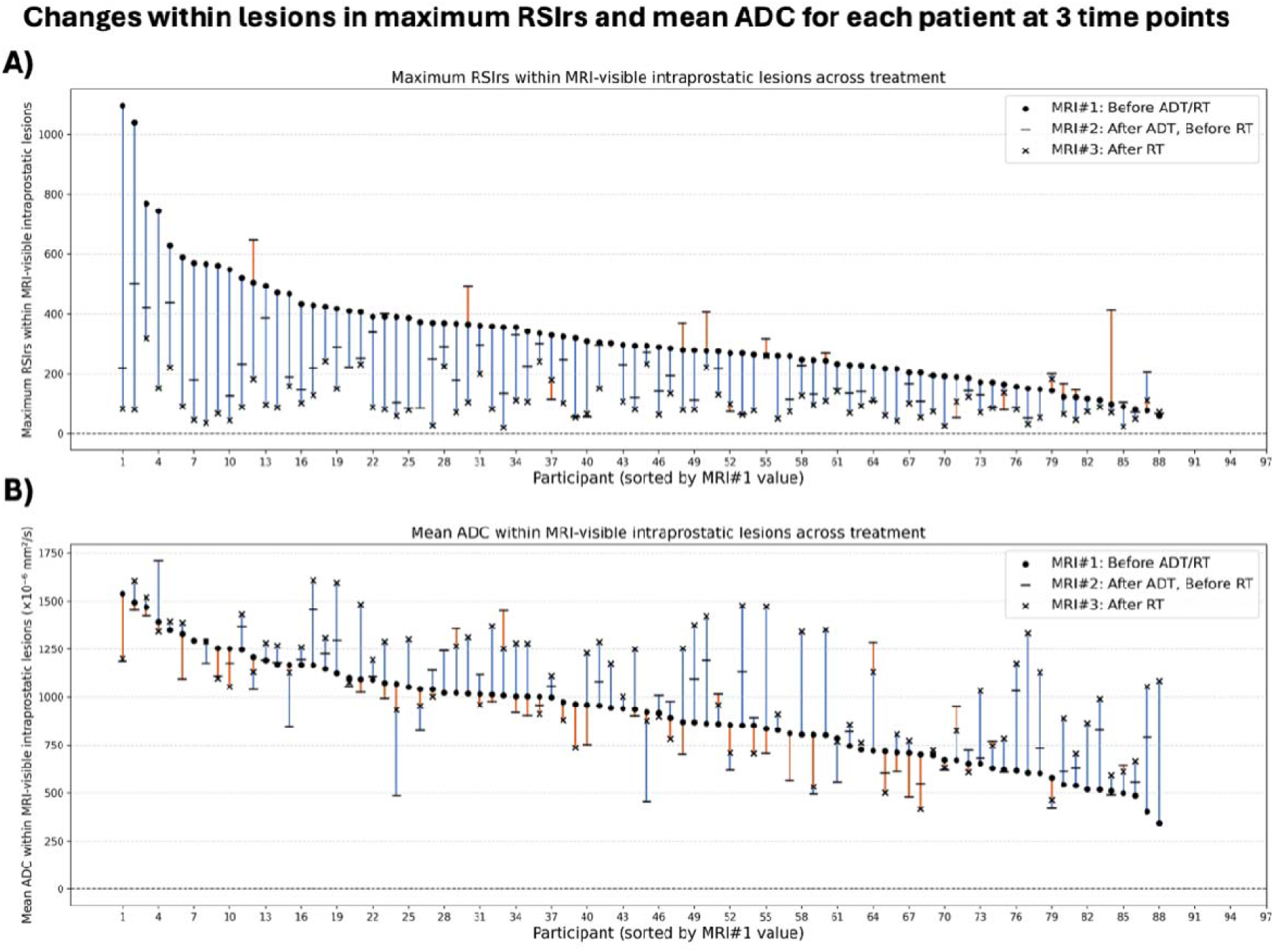
Individual changes in maximum Restriction Spectrum Imaging restriction score (RSIrs) and mean apparent diffusion coefficient (ADC) within intraprostatic lesions for 88 patients who had MRI-visible lesions identified by the radiologist. MRI #1 was acquired before ADT and RT, MRI #2 after ADT but before RT, and MRI #3 was after RT. Participants are sorted from high to low maximum RSIrs and mean ADC before treatment at baseline MRI. For most patients, maximum RSIrs decreased after neoadjuvant ADT, and dropped even more after RT. The trend for changes in mean ADC was not homogenous, with some patients having higher mean ADC after treatment while others having lower ADC values.

## Discussion

We observed significant changes in the quantitative RSIrs imaging biomarker within intraprostatic lesions following treatment, suggesting sensitivity of imaging to biological response to treatment. Observed decreases in RSIrs with treatment are generally concordant with expectations of high treatment efficacy in this population. ADC changes were less harmonious, consistent with a prior prospective study that concluded ADC and conventional DWI are not suitable for assessing treatment response^18^. While nearly all patients experienced a decrease in RSIrs within the lesion, the magnitude of the decrease varied across patients, possibly reflecting inter-patient heterogeneity in tumor response. The clinical significance of response on RSIrs will be evaluated as clinical follow-up for ARIEL matures.

Patients with PC treated with RT experience changes in gland and tumor volumes, but the pattern and extent of shrinkage are not fully understood. ADT further alters prostate volume, with studies showing reductions of 20–50% after a few months of treatment^19^. Prostate and lesion changes with ADT could affect the accuracy of treatment planning for focal RT boost. It is well known from ultrasound studies that prostate volume can decrease with ADT, though serial measurements with more precise MRI have been less common^20^. Changes in lesion visibility and size are not well described. ARIEL participants had highly variable decreases in prostate size with neoadjuvant ADT, and most had further prostate shrinkage during RT, which could be from the RT itself, ongoing ADT, or both. Either way, neoadjuvant ADT does not cause the prostate size to reach a new (smaller) steady state prior to RT. Meanwhile, 40% of visible lesions prior to treatment were no longer clearly visible after neoadjuvant ADT. Those lesions that were still visible had shrunk, on average, more than twice as much as the prostate. Given that ADT is not considered a curative therapy, these changes to the lesion may represent a loss of visibility out of proportion to actual cancer cell death. Focal boost based on post-ADT imaging may therefore lead to relative underdosing of the tumor.

Pragmatic steps are warranted to ensure accurate PC treatment in light of unpredictable prostate and lesion changes after neoadjuvant ADT. For patients that start ADT before RT, a new prostate MRI before starting RT is useful for precise delineation of the prostate. CT-based prostate contours are known to be unreliable^21^. As for lesions, given their dramatic changes— and frequent disappearance—observed in ARIEL, if focal boost is planned, pre-treatment MRI is necessary for accurate targeting. Cognitive or deformable automated registration can be used to attempt to account for changes to the lesion, though the accuracy of these is not well established. Alternatively, the ARIEL results suggest an appealing strategy is to start ADT and RT simultaneously (i.e., omit neoadjuvant ADT). Treatment is then based on only one MRI that can be used to delineate both the prostate and the focal boost target(s). This strategy spares the patient an extra MRI. And studies of ADT efficacy have suggested the adjuvant period is more important than neoadjuvant ADT^1,22^.

Prostate auto-segmentations using our validated AI model were accurate. On central radiology review, all auto-segmentations of the prostate were deemed acceptable (no major revisions required) at all time points. Minor corrections were limited to anatomically challenging regions, primarily at the prostate base and apex, which are known to show greater inter-reader variability even among expert radiologists^23^. Accurate prostate auto-segmentation on MRI in the post-treatment setting (e.g., after radiotherapy or androgen deprivation therapy) has not been systematically validated, as treatment-related changes in gland size, signal intensity, and anatomy can complicate contouring. This type of tool could possibly improve efficiency in radiation oncology workflows and help reduce variability in prostate contouring, especially among oncologists less experienced with prostate MRI.

Limitations of this prospective trial include that it was conducted at a single center. Central radiology interpretation reduces variability but cannot account for inter-reader differences. Patient-specific differences and scheduling logistics created heterogeneity in the timing of the imaging exams. Ultrahypofractionation was excluded from this trial due to concerns that its distinct impact on the timing of treatment changes and unique effects on microvasculature could be too different from longer courses of RT.

## Conclusion

ARIEL is a phase II observational trial that has completed accrual. Primary results will be presented when the primary endpoint is reached. Early results demonstrate significant changes in RSIrs for visible lesions after ADT and RT, which might possibly be indicative of tumor response. When neoadjuvant ADT is given, both pre-ADT and post-ADT MRI are likely necessary for accurate focal RT boost targeting. Concurrent commencement of ADT and RT would make focal RT boost planning less complex.

## Data Availability

All data produced in the present study are available upon reasonable request to the authors

## Funding

This work was supported, in part, by the American Society for Radiation Oncology and the Prostate Cancer Foundation (ASTRO-PCF #PCF20YOUN01), as well as the National Institutes of Health (NIH/NIBIB K08EB026503; UL1TR000100).

## Notes

### Competing Interest Statement

Dr. Tyler Seibert reports honoraria or consulting fees from Varian Medical Systems, WebMD, MJH Life Sciences, MD Education USA, GE Healthcare, Blue Earth Diagnostics, Janssen, CorTechs Labs, and MyOme. He has stock options in CorTechs Labs, MyOme, and Open Medicine for serving on their scientific advisory boards. He receives research funding and/or in-kind research support from GE Healthcare, Blue Earth Diagnostics, Quibim, AIRA Matrix, Veracyte, and Lantheus, all through the University of California San Diego. These companies might potentially benefit from the research results. The terms of this arrangement have been reviewed and approved by the University of California San Diego in accordance with its conflict-of-interest policies.
Dr. Anders Dale is a Founding Director and holds equity in CorTechs Labs, Inc. (DBA Cortechs.ai), Precision Pro, Inc., and Precision Health and Wellness, Inc. AMD is the President and a Board of Trustees member of the J. Craig Venter Institute (JCVI). He is a Professor II at Oslo University in Norway.

### Clinical Trial

NCT04349501

### Author Declarations

Ethics committee/IRB of UC San Diego gave ethical approval for this work

## References

1. Spratt DE, Srinivas S, Adra N, et al. Prostate Cancer, Version 3.2026, NCCN Clinical Practice Guidelines In Oncology. J Natl Compr Canc Netw. 2025;23(11):469–493. doi:10.6004/jnccn.2025.0052

2. Conlin CC, Feng CH, Digma LA, et al. A Multicompartmental Diffusion Model for Improved Assessment of Whole-Body Diffusion-weighted Imaging Data and Evaluation of Prostate Cancer Bone Metastases. Radiology: Imaging Cancer. 2023;5(1):e210115. doi:10.1148/rycan.210115

3. Zhong AY, Digma LA, Hussain T, et al. Automated Patient-level Prostate Cancer Detection with Quantitative Diffusion Magnetic Resonance Imaging. European Urology Open Science. 2023;47:20–28. doi:10.1016/j.euros.2022.11.009

4. Feng CH, Conlin CC, Batra K, et al. Voxel-level Classification of Prostate Cancer on Magnetic Resonance Imaging: Improving Accuracy Using FOUR-COMPARTMENT Restriction Spectrum Imaging. Magnetic Resonance Imaging. 2021;54(3):975–984. doi:10.1002/jmri.27623

5. Domingo MR, Do DD, Conlin CC, et al. Restriction Spectrum Imaging as a quantitative biomarker for prostate cancer with reliable positive predictive value. The Journal of Urology. Published online November 20, 2024. doi:10.1097/JU.0000000000004611

6. Kallis K, Conlin CC, Ollison C, et al. Quantitative MRI biomarker for classification of clinically significant prostate cancer: Calibration for reproducibility across echo times. J Appl Clin Med Phys. 2024;25(11):e14514. doi:10.1002/acm2.14514

7. Kallis K, Conlin CC, Zhong AY, et al. Comparison of synthesized and acquired high b-value diffusion-weighted MRI for detection of prostate cancer. Cancer Imaging. 2024;24:89. doi:10.1186/s40644-024-00723-6

8. Do DD, Rojo Domingo M, Conlin CC, et al. Robustness of a Restriction Spectrum Imaging (RSI) quantitative MRI biomarker for prostate cancer: assessing for systematic bias due to age, race, ethnicity, prostate volume, medication use, or imaging acquisition parameters. Preprint posted online September 12, 2024. doi:10.1101/2024.09.10.24313042

9. Kerkmeijer LGW, Groen VH, Pos FJ, et al. Focal Boost to the Intraprostatic Tumor in External Beam Radiotherapy for Patients With Localized Prostate Cancer: Results From the FLAME Randomized Phase III Trial. J Clin Oncol. 2021;39(7):787–796. doi:10.1200/JCO.20.02873

10. Zhong AY, Lui AJ, Katz MS, et al. Use of focal radiotherapy boost for prostate cancer: radiation oncologists’ perspectives and perceived barriers to implementation. Radiat Oncol. 2023;18(1):188. doi:10.1186/s13014-023-02375-5

11. Dornisch AM, Zhong AY, Poon DMC, Tree AC, Seibert TM. Focal radiotherapy boost to MR-visible tumor for prostate cancer: a systematic review. World J Urol. 2024;42(1):56. doi:10.1007/s00345-023-04745-w

12. Rojo Domingo M, Conlin CC, Karunamuni R, et al. Utility of quantitative measurement of T2 using restriction spectrum imaging for detection of clinically significant prostate cancer. Sci Rep. 2024;14(1):31318. doi:10.1038/s41598-024-82742-8

13. Ko EC, Stone NN, Stock RG. PSA nadir of <0.5 ng/mL following brachytherapy for early-stage prostate adenocarcinoma is associated with freedom from prostate-specific antigen failure. Int J Radiat Oncol Biol Phys. 2012;83(2):600–607. doi:10.1016/j.ijrobp.2011.07.009

14. Song Y, Dornisch AM, Dess RT, et al. Multidisciplinary Consensus Prostate Contours on Magnetic Resonance Imaging: Educational Atlas and Reference Standard for Artificial Intelligence Benchmarking. International Journal of Radiation Oncology*Biology*Physics. Published online March 2025. doi:10.1016/j.ijrobp.2025.03.024

15. Nguyen L, Song Y, Dornisch A, et al. PURE-MRI: An international study assessing physician accuracy in delineating the prostate and urethra on prostate MRI. Radiotherapy and Oncology. 2026;216. doi:10.1016/j.radonc.2025.111333

16. Song Y, Nguyen L, Dornisch A, et al. Prostate Auto-Segmentation Model for MRI with or without Hydrogel Spacer. International Journal of Radiation Oncology, Biology, Physics. 2025;123(1):e608–e609. doi:10.1016/j.ijrobp.2025.06.2879

17. Scott R, Misser SK, Cioni D, Neri E. PI-RADS v2.1: What has changed and how to report. SA J Radiol. 2021;25(1):2062. doi:10.4102/sajr.v25i1.2062

18. Barrett T, Gill AB, Kataoka MY, et al. DCE and DW MRI in monitoring response to androgen deprivation therapy in patients with prostate cancer: A feasibility study. Magnetic Resonance in Medicine. 2012;67(3):778–785. doi:10.1002/mrm.23062

19. Sanguineti G, Marcenaro M, Franzone P, Foppiano F, Vitale V. Neoadjuvant androgen deprivation and prostate gland shrinkage during conformal radiotherapy. Radiotherapy and Oncology. 2003;66(2):151–157. doi:10.1016/S0167-8140(03)00031-8

20. Hötker AM, Mazaheri Y, Zheng J, et al. Prostate Cancer: Assessing the effects of androgen-deprivation therapy using quantitative multi-parametric MRI. Eur Radiol. 2015;25(9):2665–2672. doi:10.1007/s00330-015-3688-1

21. McLaughlin PW, Evans C, Feng M, Narayana V. Radiographic and anatomic basis for prostate contouring errors and methods to improve prostate contouring accuracy. Int J Radiat Oncol Biol Phys. 2010;76(2):369–378. doi:10.1016/j.ijrobp.2009.02.019

22. Spratt DE, Malone S, Roy S, et al. Prostate Radiotherapy With Adjuvant Androgen Deprivation Therapy (ADT) Improves Metastasis-Free Survival Compared to Neoadjuvant ADT: An Individual Patient Meta-Analysis. J Clin Oncol. 2021;39(2):136–144. doi:10.1200/JCO.20.02438

23. Song Y, Dornisch A, Dess RT, et al. Precise prostate contours: setting the bar and meticulously evaluating AI performance. Preprint posted online October 22, 2024. doi:10.1101/2024.10.21.24315771

